# GFAP as a Marker of Neuroinflammation in Patients with COVID-19: A Comparative Analysis with Alzheimer’s and Parkinson’s Disease

**DOI:** 10.1101/2025.09.12.25335574

**Authors:** Oleg S. Popov, Natalya N. Sushentseva, Svetlana V. Apalko, Anna Yu. Asinovskaya, Sergey V. Mosenko, Andrey M. Sarana, Sergey G. Shcherbak

## Abstract

Background: Persistent neurocognitive symptoms following COVID-19 may stem from sustained neuroinflammation. Glial fibrillary acidic protein (GFAP) is a blood marker of astroglial activation; how it aligns COVID-19 with neurodegenerative diseases remains unclear. Methods: We compared plasma GFAP and a focused panel (fractalkine/CX3CL1, MDC/CCL22, sCD40L, IP-10/CXCL10, VEGFA) across patients hospitalized with COVID-19 who later developed post-COVID neurological symptoms (n=36), Alzheimer’s disease (AD, n=40), Parkinson’s disease (PD, n=44), and controls (n=30). Assays used xMAP Luminex (panel) and ELISA (GFAP). Non-parametric statistics (Kruskal–Wallis; Mann–Whitney, α=0.01) and multivariate analyses (PCA, LDA) were performed on log-transformed, standardized data in R 4.5.1. Results: GFAP was elevated versus controls in COVID-19 (median 308 pg/mL) and PD (475 pg/mL) and modestly increased in AD (250 pg/mL) compared with controls (194 pg/mL). COVID-19 showed a distinct inflammatory signature with markedly reduced fractalkine and MDC, and increased IP-10, while AD/PD exhibited higher sCD40L and VEGFA. GFAP did not differ between COVID-19 and AD, positioning astroglial activation as a shared axis. PCA/LDA placed COVID-19 between controls and AD/PD, indicating partial convergence with neurodegeneration yet a unique acute-inflammation profile. Conclusions: Blood GFAP aligns COVID-19 with AD/PD on astrocytic activation, whereas chemokine and platelet-activation markers distinguish COVID-19 from chronic neurodegeneration. These findings support composite biomarker panels for stratifying post-COVID neuroinflammation.

## Introduction

A substantial subset of individuals recovering from COVID-19 experience persistent fatigue, cognitive inefficiency, and other neurological symptoms [1,2]. Chronic neuroinflammation—potentially initiated by systemic cytokine surges and blood–brain barrier dysfunction—has been proposed as a key mechanism. Because astrocytes respond to CNS injury and inflammation, plasma GFAP has emerged as an accessible marker of reactive astrogliosis [3]. However, it remains important to delineate whether the COVID-19 biomarker profile merely mirrors chronic neurodegeneration or carries distinguishing features [4]. We therefore quantified GFAP alongside fractalkine (CX3CL1), MDC/CCL22, soluble CD40 ligand (sCD40L), IP-10/CXCL10, and VEGFA, and compared profiles in COVID-19 versus AD, PD, and controls.

## Methods

Design and cohorts: We analyzed banked serum from four groups: hospitalized COVID-19 patients (PCR-confirmed, April 2020–March 2022) who were later identified (November 2022–January 2024) with post-COVID neurological symptoms (n=36); AD (n=40) diagnosed per NIA criteria with FDG-PET support; PD (n=44); and controls without neurological symptoms (n=30) (table 1).

**Table 1.** Sex and age composition of the main study groups stratified by outcomes and disease severity (mean ± SD). COVID-19 — patients diagnosed with “coronavirus disease COVID-19” (U07.1); AD — patients diagnosed with “Alzheimer’s disease” (G30); PD — patients diagnosed with “Parkinson’s disease” (G20); Control — apparently healthy participants.

| Group | Total (n) | Male, n | Female, n | Age, overall (years) | Age, males (years) | Age, females (years) |
| --- | --- | --- | --- | --- | --- | --- |
| COVID-19 | 36 | 14 | 22 | 53.2±7.3 | 50.9±6 | 54.7±7.8 |
| AD | 40 | 9 | 31 | 68.7±5.8 | 68.4±5.3 | 68.8±6 |
| PD | 44 | 16 | 28 | 61.6±8 | 57.9±9.5 | 63.6±6.3 |
| Control | 30 | 14 | 16 | 54.6±7 | 53±6.6 | 56±7.3 |

Infection markers and therapy were not initiated before sampling. Key exclusions included HIV, syphilis, and viral hepatitis. Biomarker assays: sCD40L, MDC, fractalkine, IP-10, and VEGFA were measured by xMAP Luminex; GFAP by ELISA (Abbexa). Statistics: Missing values were imputed by MCMC with predictive mean matching after diagnostics for convergence. Normality was screened by Shapiro–Wilk. Group differences used Kruskal–Wallis with Mann–Whitney post-hoc tests (α=0.01). Multivariate structure was explored via PCA and LDA on log-transformed, standardized concentrations (R 4.5.1). Ethics: The study was approved by the Ethics Committee of City Hospital No. 40 (protocol number 215, April 20, 2022), with written informed consent; not a clinical trial.

## Results

Univariate differences: All six biomarkers differed across the four groups (Kruskal– Wallis p<0.01). GFAP was higher in COVID-19 and PD versus controls (e.g., COVID-19 median 308 pg/mL vs controls 194 pg/mL; AD median 250 pg/mL; PD median 475 pg/mL), with COVID-19 > controls at p=0.002 (Figure 1).

**Figure 1.**
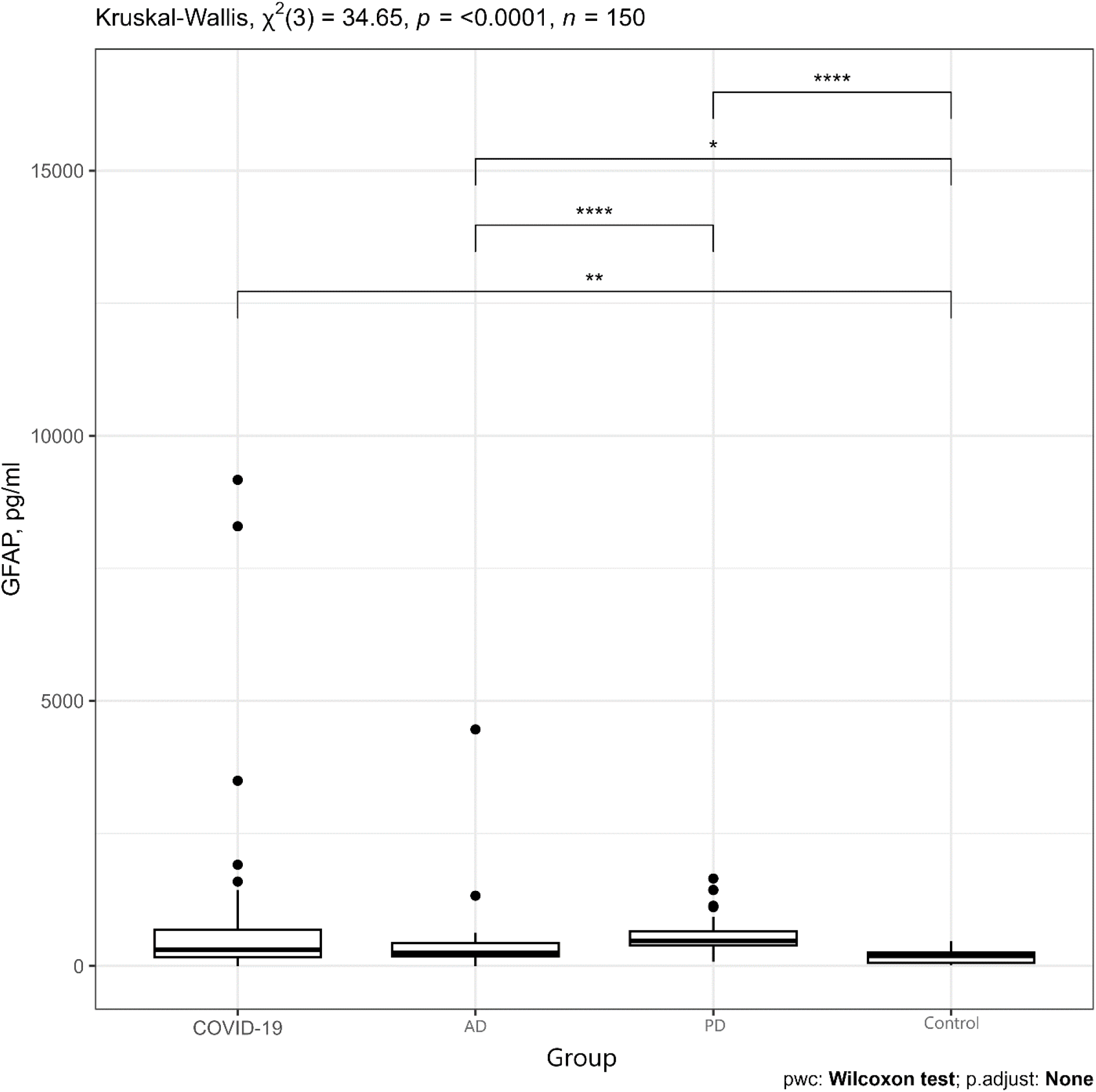
Box-and-whisker plot of GFAP concentration with the Kruskal–Wallis test result and pairwise post hoc comparisons for each group (Mann–Whitney U test). COVID-19 — patients diagnosed with “coronavirus disease COVID-19” (U07.1); AD — patients diagnosed with “Alzheimer’s disease” (G30); PD — patients diagnosed with “Parkinson’s disease” (G20); Control — apparently healthy participants. Asterisks denote significance levels: ^*^ p < 0.05; ^**^ p < 0.01; ^****^ p < 0.001.

COVID-19 showed sharply reduced fractalkine and lower MDC compared with other groups (e.g., COVID-19 MDC median 375 pg/mL vs controls 769 pg/mL, p=0.002), while IP-10 was markedly increased in COVID-19 but lowest in AD (Figure 2).

**Figure 2.**
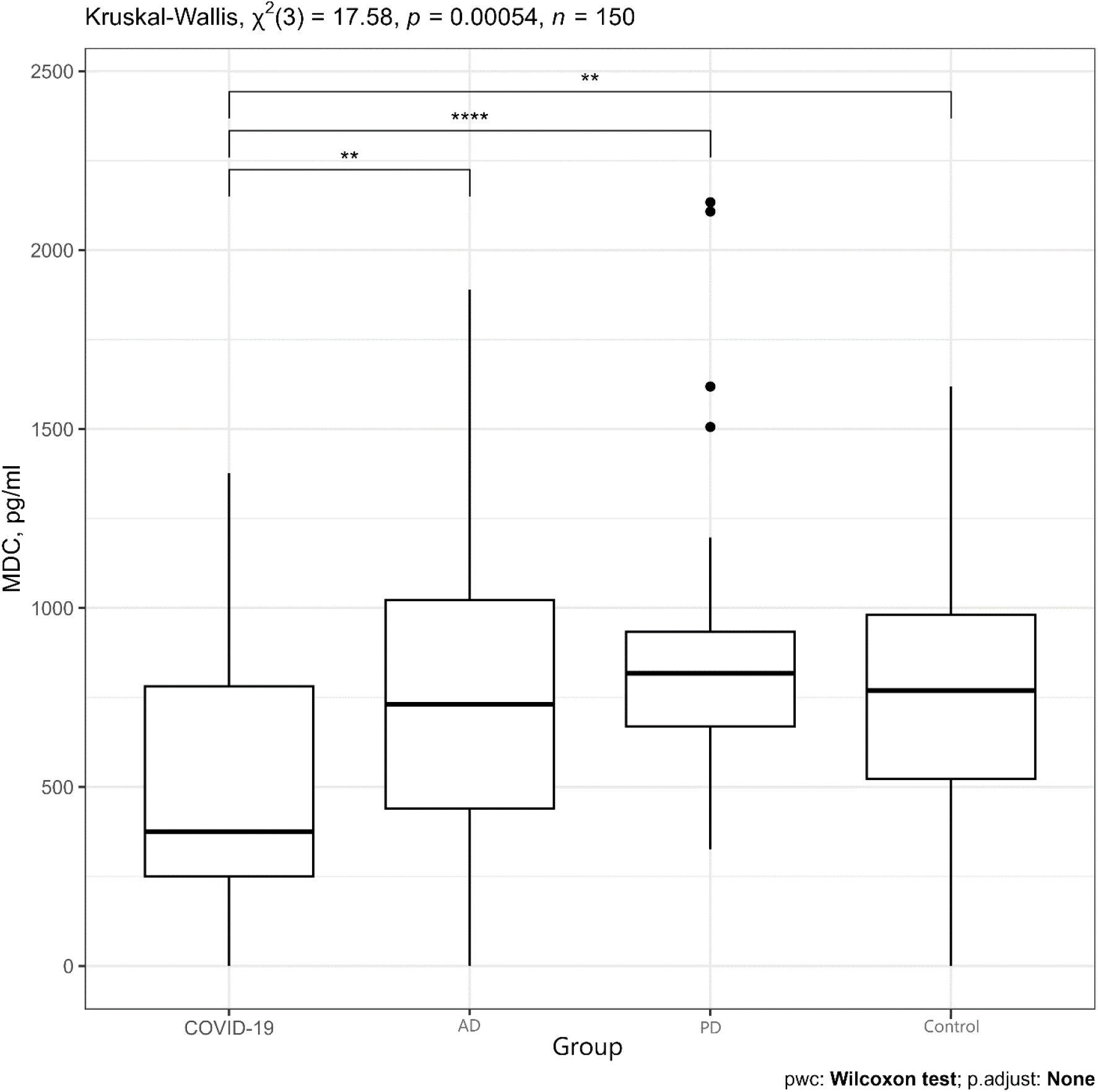
Box-and-whisker plot of MDC concentration with the Kruskal–Wallis test result and pairwise post hoc comparisons for each group (Mann–Whitney U test). COVID-19 — patients diagnosed with “coronavirus disease COVID-19” (U07.1); AD — patients diagnosed with “Alzheimer’s disease” (G30); PD — patients diagnosed with “Parkinson’s disease” (G20); Control — apparently healthy participants. Asterisks denote significance levels: ^*^ p < 0.05; ^**^ p < 0.01; ^****^ p < 0.001.

sCD40L and VEGFA were highest in AD/PD; VEGFA exceeded controls in both neurodegenerative cohorts (p<0.001). Multivariate patterning: PCA and LDA placed controls far from patient groups and positioned COVID-19 between controls and AD/PD. GFAP contributed most strongly to patient–control separation, and was the only marker without a significant difference between COVID-19 and AD, reinforcing astroglial activation as a shared feature. By contrast, chemokine and platelet-activation signals (low fractalkine/MDC, high IP-10; high sCD40L/VEGFA in AD/PD) differentiated COVID-19 from chronic neurodegeneration.

## Discussion

Our findings indicate partial convergence of COVID-19 with AD/PD via astrocytic activation (elevated GFAP), alongside a distinct COVID-19 signature consistent with acute systemic inflammation. This duality explains why COVID-19 clusters nearer to neurodegenerative cohorts than to controls in multivariate space, yet remains separable by chemokine and platelet-activation markers [5]. Clinically, composite panels centered on GFAP may help stratify post-COVID patients at risk for persistent neurological symptoms. Limitations include single-center sampling, moderate cohort sizes, and cross-sectional design without longitudinal trajectories; causal inferences cannot be drawn.

## Data Availability

All data produced in the present study are available upon reasonable request to the authors

## Declarations

### Ethics approval and consent to participate

Approved by the Ethics Committee of City Hospital No. 40, protocol number 215 (April 20, 2022); written informed consent obtained.

### Consent for publication

Not applicable (de-identified data).

### Availability of data and materials

De-identified data and analysis code are available from the corresponding author upon reasonable request.

### Authors’ contributions

Concept and design: S.G. Shcherbak, A.M. Sarana, S.V. Apalko. Material collection and processing: A.Yu. Asinovskaya, S.V. Mosenko, N.N. Sushentseva. Statistical analysis: O.S. Popov. Manuscript drafting: O.S. Popov, N.N. Sushentseva. Critical revision: S.V. Apalko, O.S. Popov, N.N. Sushentseva. All authors approved the final manuscript.

### License

Suggested for medRxiv: CC BY 4.0 (authors may adjust on submission).

